# Increased dose of H1N1 pandemic influenza vaccine during pregnancy improves immunity in mothers and infants

**DOI:** 10.1101/2025.11.18.25340497

**Authors:** Dhvanir Kansara, Martina Kosikova, Patricia L. Milletich, Jie Zhou, Lynda Coughlan, Michael S Zens, Geeta Swamy, Anne G. Hoen, Hang Xie, Margaret E. Ackerman, Marcela F. Pasetti, the DMID 09-0072 Clinical Study Group

## Abstract

Maternal-infant immunity against influenza is improved through vaccination during pregnancy. We conducted an in-depth analysis of antibody (Ab) responses in sera from pregnant and non-pregnant women immunized with an unadjuvanted inactivated influenza A (H1N1) monovalent vaccine during the 2009 pandemic (NCT00992719). Pregnant women received either the standard 15µg- or increased 30µg-dose, while non-pregnant women received the 15µg-dose. Ab specific for influenza hemagglutinin (HA), HA stalk, and neuraminidase (NA), as well as canonical functions of hemagglutination inhibition (HAI), microneutralization, and neuraminidase inhibition were examined at baseline, 21 days post-vaccination, at delivery, and in cord blood. Ab subclasses, Fc receptor binding, and Fc-mediated immune functions, including cellular cytotoxicity, phagocytosis, and complement deposition, were also assessed. The vaccine was well-tolerated and highly immunogenic in recipients; most participants had a ≥4-fold increase in Ab titers post-vaccination for HAI (HAI> 70%) and HA-specific IgG (IgG> 50%). Pregnant women who received the 15µg dose had a lower vaccine response in terms of NA-specific IgG and Fc receptor binding compared to the other groups. Immunization of pregnant women with the 30µg-dose resulted in more robust humoral immunity, including a larger number of HA Ab features reaching 4-fold increases compared to the other groups and a more durable antiviral function, and increased NA-specific Ab features that were transferred to the infant as compared to pregnant women who received the standard 15µg-dose. Increasing the antigen content in seasonal vaccines could be a means to enhance immunity against influenza in mothers and infants and deserves further study.

**Importance:** Pregnant women and infants are at-risk groups for influenza infection. Vaccination is recommended during pregnancy to stimulate adaptive immunity and protect both the mother and infant early in life. We characterized the immune responses of pregnant and non-pregnant women to an unadjuvanted inactivated 2009 pandemic influenza A (H1N1). Pregnant women immunized with the 15µg standard seasonal influenza vaccine dose developed lower NA-specific responses compared to non-pregnant women immunized with the same vaccine dose. Vaccination of pregnant women with an increased 30µg dose resulted in more robust and durable responses post-vaccination, particularly longer-lasting functional antibodies and NA-specific antibodies in maternal and cord blood. A deeper analysis of antibody responses beyond the traditional hemagglutination inhibition (HAI), suggests that a higher-dose influenza vaccine, already recommended for the elderly, could be beneficial for pregnant women and is worth exploring.

## Introduction

Pregnant and postpartum women are at increased risk for severe illness and serious complications associated with influenza infection (1, 2). This higher risk for disease was exemplified during the 2009 H1N1 pandemic, wherein pregnant and postpartum women had a disproportionately higher rate of hospital admission and influenza-associated death as compared to the general population (3–5). Children of affected women were more likely to be born pre-term and to experience adverse outcomes (6); children also had the highest rate of hospitalization and death due to influenza infection as compared to other age groups (7).

Maternal vaccination is an effective intervention to prevent influenza and influenza-related complications in the mother-infant dyad and is recommended at any stage of pregnancy during the flu season (8, 9). Importantly, passively acquired maternal immunity reduces the infant’s susceptibility during the first months of life. Closing the gap of early life immunity is necessary, as seasonal influenza vaccines are only approved for infants six months of age and older.

On September 15, 2009, in the wake of the swine flu pandemic, the United States Food and Drug Administration (US FDA) approved and licensed an inactivated H1N1 vaccine manufactured by Novartis, which was recommended for healthy individuals ≥4 years of age (10). The Centers for Disease Control and Prevention (CDC) Advisory Committee on Immunization Practices (ACIP) ranked pregnant women in the highest priority group for vaccination (11). Based on experience with traditional seasonal vaccines and emerging 2009 H1N1 data, it was suggested that the standard 15μg dose of this vaccine would be immunogenic for expectant mothers and other groups with increased risk (12).

However, unlike other physiological conditions, pregnancy involves distinct stages of unique tolerogenic immunological adaptations to support fetal growth and full-term delivery (13). NK and T cell numbers and functions decrease during the second and third trimesters, signaling a shift away from inflammatory Th1 responses (14). Additionally, modulation of humoral immunity during pregnancy involves an increase in interleukin (IL)-10-secreting regulatory B cells, which may influence immune responses to vaccines (15, 16). Several studies have reported lower responses to seasonal influenza vaccination in pregnant women as compared to non-pregnant women (17, 18).

One approach to enhance vaccine-induced immunity and effectiveness, particularly for susceptible populations, is to increase the amount of antigen contained in the vaccine. Studies evaluating the effect of hemagglutinin (HA) dosage on immune responses to seasonal inactivated influenza vaccines (IIV) have demonstrated dose-related increases in serum and mucosal Ab responses (19). A high-dose IIV (60 μg of HA per strain) has been approved for adults aged 65 years or older and has been shown to be more efficacious than the standard 15 μg dose in this population (20). A higher dose vaccine may also be beneficial for pregnant women.

In the wake of the 2009 H1N1 pandemic (H1N1 pdm09), a Phase II clinical study sponsored by the National Institute of Allergy and Infectious Disease (NIAID) Division of Microbiology and Infectious Diseases (DMID, protocol 09-0072, NCT00992719) was conducted to investigate the safety and immunogenicity of the Novartis unadjuvanted H1N1 pdm09 IIV administered to pregnant women at two different dosage levels: 15μg and 30μg, and to a non-pregnant control group at the standard 15μg-dose (10, 21). Here, we describe a high-dimensional analysis of vaccine responses and report detailed features of humoral immunity at baseline, 21 days post-vaccination, at the time of delivery, and in cord blood. The magnitude and functional capabilities of vaccine-induced antibodies, as well as their durability and transfer to cord blood, were examined and compared among treatment groups to determine the impact of pregnancy status and vaccine dose on influenza immunity.

## Results

### Study design and vaccine tolerability

We conducted an in-depth analysis of Ab responses in serum samples from an open-label Phase II clinical study that investigated the safety, reactogenicity, and immunogenicity of the Novartis H1N1 pdm09 monovalent IIV administered intramuscularly to pregnant and non-pregnant women (DMID protocol 09-0072; NCT00992719). Pregnant women were randomized to receive a 15μg dose of the H1N1 pdm09 IIV vaccine (P15, n=28) or a 30μg dose (P30, n=28), while a non-pregnant control group received a 15μg dose (NP15, n=28), which is standard for seasonal influenza vaccination.

The vaccine was well tolerated **(Table 1)**; no differences were observed in the number of participants reporting solicited local reactions after vaccination, either subjective (pain, tenderness, or swelling) or quantitative events (redness or swelling measured in millimeters) among the different treatment groups (chi-sq p > 0.05). Systemic reactions (fever, malaise, myalgia, and nausea) were comparable among the cohorts, except for malaise, which was higher in the P30 group (chi-sq p=0.01). No vaccine-associated serious adverse events were reported for any treatment group. No differences were found in gestational age at vaccination or in the interval between immunization and delivery among the pregnant women’s groups (**Table 1**). Additionally, no differences were seen in the reporting of maternal complications during pregnancy, labor, and delivery, or in neonatal complications between the pregnant groups (21).

**Table 1.** Description of adverse events and cohorts.

|  | Non-pregnant<br>standard dose<br>15µg HA | Pregnant<br>standard dose<br>15µg HA | Pregnant<br>high dose<br>30µg HA | p value |
| --- | --- | --- | --- | --- |
| **Number of Participants**<br>**Vaccinated** | 28 | 28 | 28 |  |
| **Solicited Local Reactions After Vaccination N(%)** |  |  |  |  |
| Pain | 6 (21.4%) | 4 (14.3%) | 8 (28.6%) | 0.43<sup>B</sup> |
| Tenderness | 14 (50.0%) | 13 (46.4%) | 15 (53.6%) | 0.87<sup>B</sup> |
| Swelling | 2 (7.1%) | 1 (3.6%) | 2 (7.1%) | 0.81<sup>B</sup> |
| Redness (Quantitative) | 2 (7.1%) | 2 (7.1%) | 2 (7.1%) | 1.00<sup>B</sup> |
| Swelling (Quantitative) | 2 (7.1%) | 1 (3.6%) | 1 (3.6%) | 0.77<sup>B</sup> |
| **Solicited Systemic Reactions After Vaccination N(%)** |  |  |  |  |
| Feverish | 3 (10.7%) | 0 (0.0%) | 2 (7.1%) | 0.23<sup>B</sup> |
| Malaise | 4 (14.3%) | 7 (25.0%) | 14 (50.0%) | **0.01<sup>B</sup>** |
| Myalgia | 4 (14.3%) | 1 (3.6%) | 6 (21.4%) | 0.14<sup>B</sup> |
| Headache | 7 (25.0%) | 8 (28.6%) | 9 (32.1%) | 0.84<sup>B</sup> |
| Nausea | 3 (10.7%) | 3 (10.7%) | 8 (28.6%) | 0.12<sup>B</sup> |
| **Number of Samples**<br>**Analyzed** | 23 | 21 | 23 | - |
| Median age in years<br>(IQR) | 30<br>(24.5, 35) | 31<br>(27, 35) | 28<br>(25.5, 33.5) | 0.70<sup>A</sup> |
| **Race N(%)** |  |  |  |  |
| Asian | 2 (8.7%) | 2 (9.5%) | 4 (17.4%) | 0.87<sup>B</sup> |
| Black or African American | 4 (17.4%) | 3 (14.3%) | 5 (21.7%) |  |
| Multiple race | 1 (4.3%) | 1 (4.8%) | 0 (0%) |  |
| White | 16 (70.0%) | 15 (71.4%) | 14 (60.9%) |  |
| Median gestational age at<br>vaccination in weeks (IQR) |  | 21.8<br>(17.6, 26.3) | 25.6<br>(19.9-29.6) | 0.10<sup>A</sup> |
| Median days between<br>vaccination and delivery (IQR) |  | 120<br>(85, 160) | 95<br>(64, 134) | 0.08<sup>A</sup> |
| Median gestational age at<br>delivery |  | 39<br>(39, 40) | 39<br>(37.5, 40) | 0.70<sup>A</sup> |
| **Infant sex, number (%)** |  |  |  | **<0.001** |
| Female |  | 8 (38%) | 11 (48%) |  |
| Male |  | 12 (57%) | 9 (39%) |  |
| No live birth |  | 1 (4.8%) | 0 (0%) |  |
| Multiparity |  | - | 3 (12.9%)<sup>1</sup> |  |

<sup>A</sup> Kruskal-Wallis rank sum test; Fisher's exact test
<sup>B</sup> Chi-squared test
<sup>1</sup> One set of triplets (two female, one male), two sets of twins (one set both male, one set one male, one female)

### Seroprotective vaccine responses based on traditional influenza Ab measurements

A limited analysis of serological responses using traditional assays was conducted at the time of the study and reported in ClinicalTrials.gov. Here, we present a comprehensive and detailed analysis of vaccine-induced humoral immunity using multiple target antigens and new assays. A comparative analysis of the magnitude of Ab elicited by the H1N1 pdm09 vaccine, which included canonical functional activity: hemagglutination inhibition (HAI), microneutralization (MN), and neuraminidase (NA) inhibition (NAI), as well as full-length H1 hemagglutinin (HA FL), H1 HA Stalk domain (HA Stalk), and NA-specific IgG binding at day 21 post-vaccination, demonstrated robust humoral responses across treatments (**Figure 1**). There were no differences in baseline immunity across groups, and all three arms responded to vaccination with increased levels of Ab mediating HAI, MN, and NAI as well as IgG specific for HA FL, HA Stalk, and NA **(Figure 1A)**. Mean fold changes in Ab levels over baseline and seroconversion rates, defined as the proportion of individuals with ≥4-fold increase in Ab levels post-vaccination, varied across Ab assessments **(Figure 1B)**.

**Figure 1.**
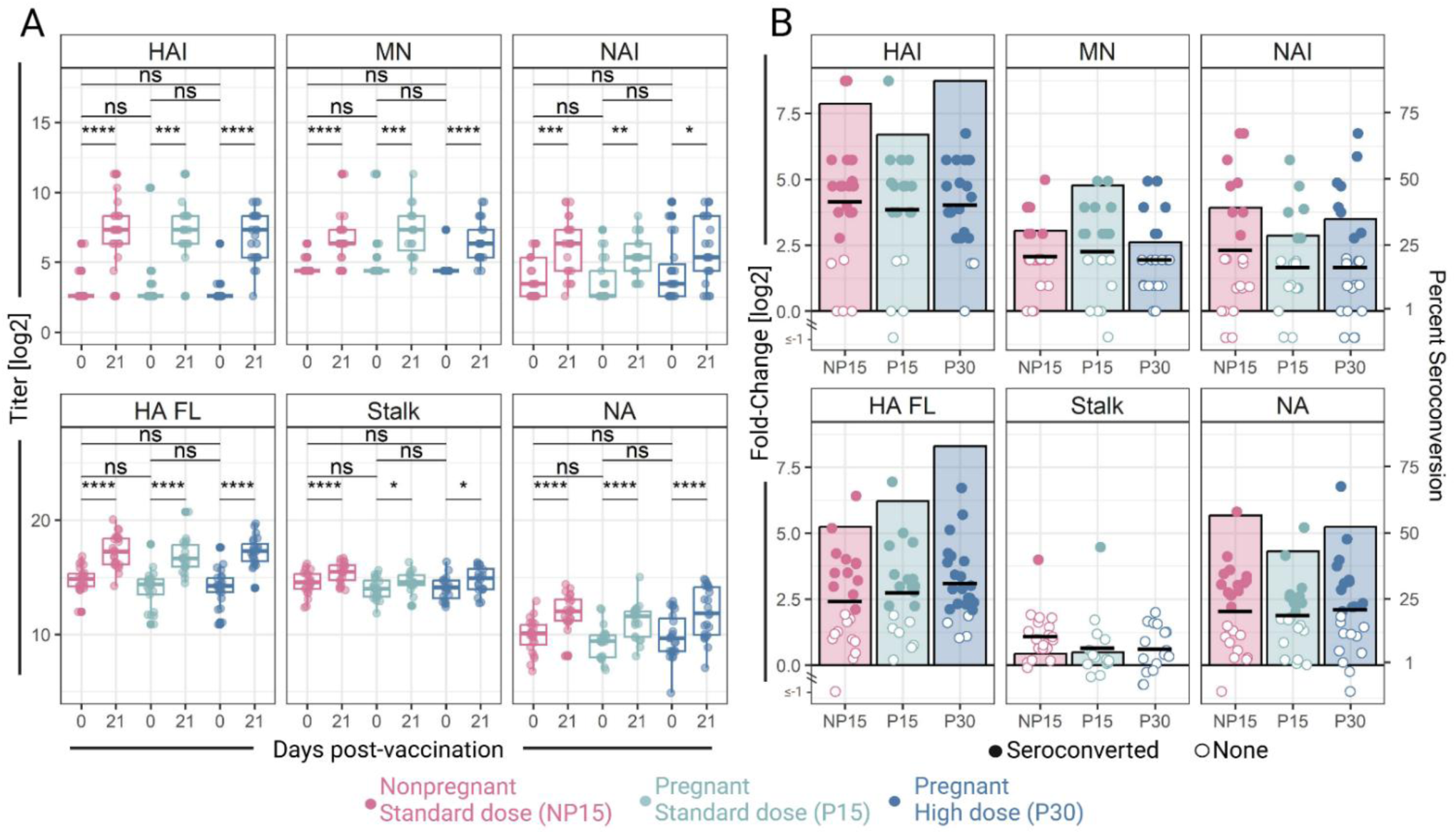
Antibody responses elicited by Novartis H1N1 pdm09 monovalent IIV in pregnant and non-pregnant women. Pregnant women received a 15μg (P15, n=21) or 30μg dose (P30, n=23), and a non-pregnant control group received a 15μg dose (NP15, n=23). Antibody responses were measured in sera before (day 0) and 21 days post-vaccination. **(A)** Hemagglutination inhibition (HAI), microneutralization (MN), and neuraminidase inhibition (NAI) activity, and IgG specific for full-length hemagglutinin (HA FL), HA stalk domain (Stalk), and neuraminidase (NA) from H1N1 pdm09 A/California/07/2009. Circles represent individual samples. ns = not significant, *p < 0.05, **p < 0.01, ***p < 0.001, ****p < 0.0001, calculated by Wilcoxon signed rank test. **(B)** Fold-change in Ab titers 21 days post-vaccination above baseline (left axis). Circles depict individual vaccine recipients and reflect their seroconversion status (Closed = seroconverted, defined as ≥4-fold increase in Ab levels over baseline. Open = no seroconversion). Bars represent the proportion of each cohort who reached seroconversion (right axis). The horizontal lines represent the mean fold change in Ab titers for each cohort.

Most participants achieved seroconversion by HAI (NP15 = 78.3% [18/23], P15 = 66.7% [14/21], P30 = 87.0% [20/23]). Although the P15 group had the lowest seroconversion rate, there was no statistically significant difference among the groups (chi-sq = 0.27). Seroprotection, defined as HAI titers ≥1:40, increased from <5% before vaccination to 82.6% [19/23], 80.1% [17/21], and 87.0% [20/23] in the NP15, P15, and P30 groups, respectively. Again, there was no statistically significant difference among the groups (chi-sq p = 0.81). The initial evaluation of HAI responses around the time the clinical study was conducted and which included all participants, showed similar results both in terms of seroconversion NP15 = 85.7% [24/28], P15 = 69.2% [18/26], and P30 = 92.9% [26/28] (chi-sq p = 0.06) and seroprotection rates: NP15 = 92.9% [26/28], P15 = 84.6% [22/26], and P30 = 96.4% [27/28] (chi-sq p = 0.28) (21). MN and NAI mean fold changes and seroconversion rates post-vaccination were somewhat lower than those for HAI **(Figure 1B),** yet still not significantly different among groups.

The majority of vaccine recipients seroconverted for HA-specific IgG (NP15 = 52.2% [12/23], P15 = 61.9% [13/21], P30=82.6% [19/23]), followed by NA-specific IgG with nearly half of the participants attained seroconversion (NP15 = 56.6% [13/23], P15 = 42.9% [9/21], P30=52.2% [12/23]) **(Figure 1B)**. A greater number of participants exhibited ≥4-fold increase in Ab responses (i.e., seroconverted) based on HAI, NAI, HA-specific IgG, and NA-specific IgG in the P30 group, as compared to the P15, although the differences were not statistically significant (chi-sq p > 0.05) **(Figure 1B)**. Intriguingly, HA stalk IgG titers increased in all groups post-vaccination **(Figure 1A)**, but seroconversion was minimal (<5%) **(Figure 1B)**.

### Robust responses to vaccination across Ab features and functions

Vaccine responses were further characterized by applying a systems serology approach to examine influenza-specific Ig isotypes (IgM, IgG, and IgA), subclasses (IgG 1-4 and IgA 1-2), and Fc-receptor binding, as well as antigen-specific Fc functions, including antibody-dependent complement deposition (ADCD) and cellular phagocytosis (ADCP), and antibody-dependent cellular cytotoxicity (ADCC) of infected cells as assessed by a cell-based FcγRIIa and FcγRIIIa ligation reporter assay.

For ease of visualization, HA- and NA-specific Ab responses in each of the treatment groups are depicted in **Figure 2A-C**, and volcano plots summarizing vaccine responses (fold changes in Ab levels 21 days post-vaccination over baseline) for all Ab features examined across groups are shown in **Figure 2D-G**. Among the various antigens interrogated, the HA-specific Ab features were the most enhanced post-vaccination, regardless of pregnancy status and vaccine dose **(Figure 2A)**, with about half of them exhibiting ≥4-fold increases (NP15 = 47.1% [8/17], P15 = 47.1% [8/17], P30 = 58.8% [10/17]) (**Figure 2D**). HA-specific Ab binding to Fc receptors had the largest increases, followed by total HA-IgG (measured by both ELISA and multiplex binding assay) (**Figure 2D)**. Robust IgG1 responses were induced in all groups, followed by IgG3 (**Figure 2A**). IgG2 increased post-vaccination but remained low compared to the other Ig subclasses. IgG4 was not induced in any of the cohorts (**Figure 2A**). Responses to HA Stalk were markedly different compared to full-length HA. HA Stalk-specific ADCD, IgG1, IgG (measured by ELISA), and FcγRIIb, and FcγRIIIa-binding Ab were increased in all groups following vaccination. The P15 group had the fewest HA stalk-specific Ab features increased after vaccination, compared to NP15 and P30 (5, 9, and 9, respectively; **Figure 2E)**.

**Figure 2.**
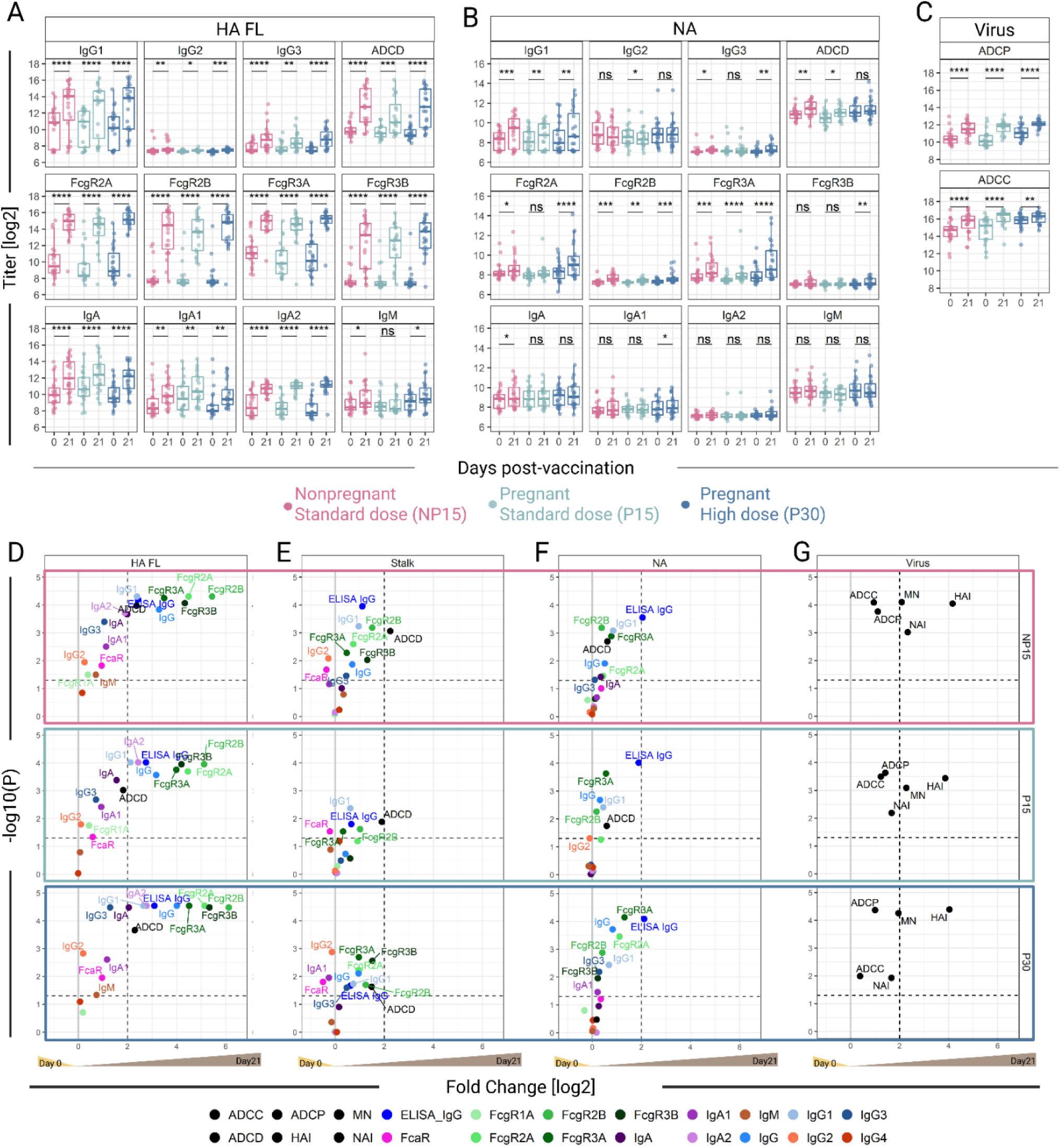
Binding and functional features of antibodies elicited by the Novartis H1N1 pdm09 IIV (A-C). Selected binding and functional features of Ab specific for **(A)** full-length hemagglutinin (HA FL) and **(B)** neuraminidase (NA), and **(C)** anti-virus activity. ns = not significant, *p < 0.05, **p < 0.01, ***p < 0.001, ****p < 0.0001, calculated by paired Wilcoxon signed-rank test. **(D-G)** Volcano plots depicting p-value (paired Wilcoxon test) on the y-axis and fold-change in Ab responses (on the x-axis) 21 days post-vaccination (Day 21) vs. baseline (Day 0) specific for **(D)** HA FL, **(E)** HA stalk domain (Stalk), **(F)** NA, and **(G)** virus. The horizontal dotted line represents p = 0.05. The vertical dotted line represents a 4-fold increase from baseline. The solid gray line represents no fold change; values above 0 had a higher Ab response post-vaccination.

NA-specific IgG, IgG1, and FcγRIIIa- and FcγRIIb-binding Ab were increased in all groups following vaccination (**Figures 2B**, **2F**). Again, the P15 group had the fewest NA-specific Ab features elevated post-vaccination as compared to NP15 and P30 (n=6, 9, and 9, respectively; **Figure 2F)**. Both NP15 and P30, but not P15, had increases in NA-specific IgG3 and FcγRIIa binding Ab (**Figures 2A and 2B**). Consistent with the robust FcγR-binding Ab responses, ADCP and ADCC were increased after vaccination in all groups (**Figure 2C)**. Among virus-blocking Ab-mediated functions, HAI had the highest mean fold increase (≥4-fold) in all treatment groups (**Figure 2G**).

Having generated both ELISA and multiplex binding data for HA, HA stalk, and NA IgG, we evaluated the agreement between the two methods. Good agreement was observed between ELISA titers and multiplex median fluorescent intensity values for all three antigens across cohorts (**Supplemental Figure 1**).

### Impact of pregnancy status and dose on vaccine responses

We next investigated differences in vaccine responses based on pregnancy status (P15 vs. NP15) and dose (P15 vs. P30) by comparing Ab responses 21 days post-vaccination (**Figure 3)**. The comparison between P30 vs. NP15 aimed to determine whether vaccination with the higher dose during pregnancy resulted in responses equivalent to those elicited by the standard dose in non-pregnant women.

**Figure 3.**
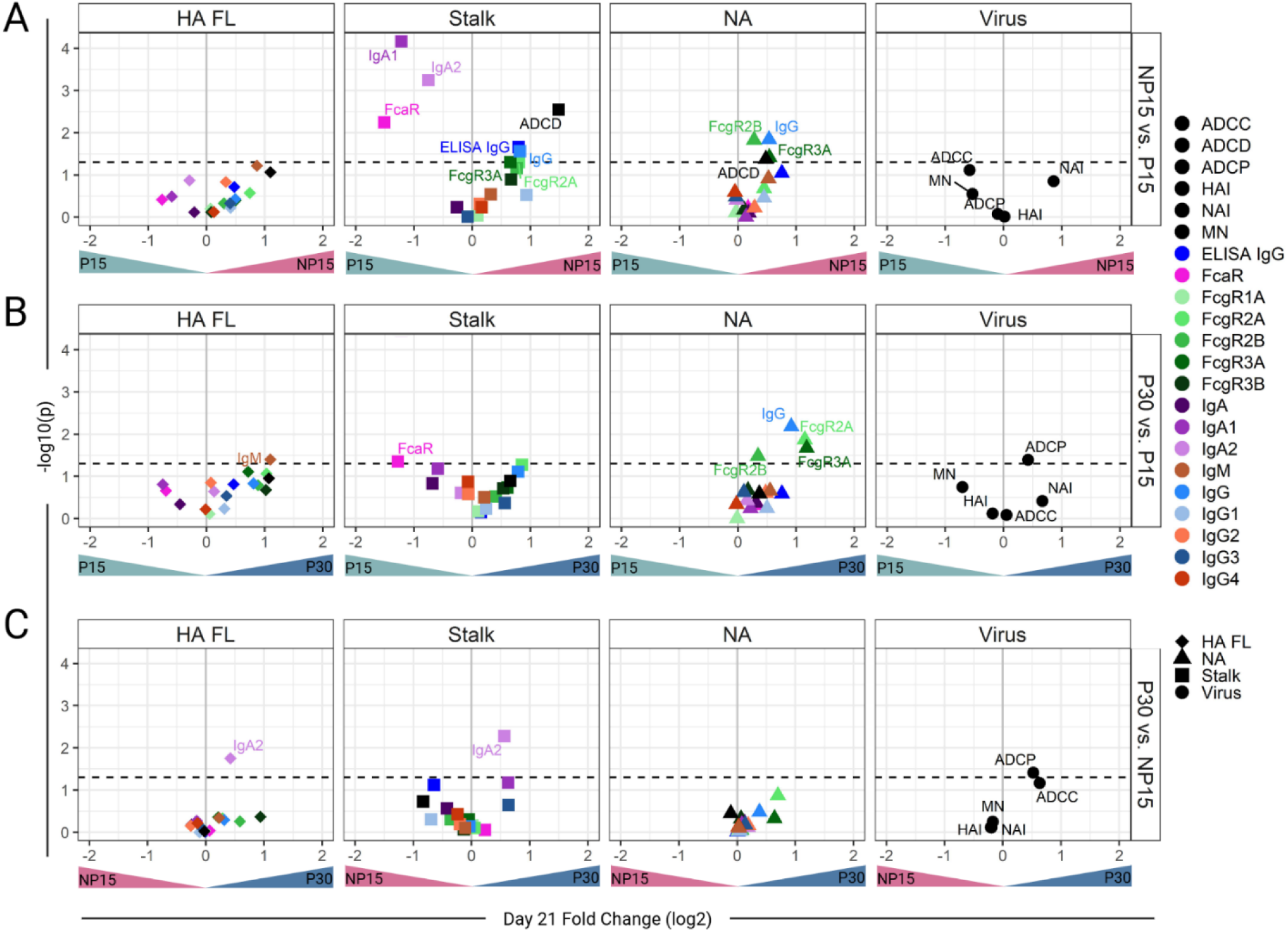
Enhanced NA antibody responses in pregnant women immunized with the standard 15μg dose of the Novartis H1N1 pdm09 IIV. Volcano plots depict p-values (paired Wilcoxon test) on the y-axis and fold-change in Ab titers 21 days post-vaccination (on the x-axis) between **(A)** pregnant women receiving 15μg (P15) vs. non-pregnant women receiving 15μg (NP15), **(B)** P15 vs. pregnant women receiving 30μg (P30), and **(C)** P30 vs. NP15. Antigen specificity (full-length hemagglutinin [HA FL], HA stalk domain [Stalk], neuraminidase [NA], and anti-virus activity are indicated by shape. Ab features are indicated by color. Horizontal dotted line represents a p-value = 0.05, and the solid grey line represents no fold-change.

Looking at the breadth of vaccine responses, represented by the number of Ab features increased post-vaccination, the P15 group had fewer Ab features increased (28.6%, 16/56), as shown in the left quadrants of **Figure 3A**, compared to the NP15 group (71.4%, 40/56). Similarly, fewer Ab features were increased in the P15 (25%, 14/56) (**Figure 3B**) as compared with the P30 vaccine recipients (75%, 42/56). Encouragingly, the P30 group had a slightly higher proportion of features elevated post-vaccination (57.1%, 32/56), as shown in the right quadrants of **Figure 3C**, compared to NP15. When considering only the Ab features that were significantly higher post-vaccination, represented by features above the horizontal dashed line, P15 still had the fewest while P30 had the most; P15<NP15 (n=3 and 9, respectively), P30>P15 (n=6 and 1, respectively), and P30>NP15 (n=3 and 0, respectively) (**Figure 3 A-C**).

HA-specific IgM and IgA2 were significantly higher in P30 as compared to P15 or NP15, respectively, while no other HA-specific feature differed among these groups. P30 also had significantly higher post-vaccination ADCP as compared to P15 or NP15. Stalk-specific Ab binding to FcαR was higher in P15 relative to NP15 and P30. Additionally, IgA1 and IgA2 were higher in P15 compared to NP15. HA-stalk-specific ADCD, IgG [measured by ELISA and multiplex binding assay], and FcγRIIIa and FcγRIIa binding were higher in NP15 compared to P15. Only HA-stalk-specific IgA2 was increased in P30 relative to NP15.

Notably, NA-specific IgG [measured by multiplex binding assay], FcγRIIb, and FcγRIIIa binding Ab were significantly increased post-vaccination, both in the NP15 and P30 groups, compared to P15. The NP15 group also had a significantly higher NA-specific ADCD than the P15 group. P30 exhibited higher NA-specific FcγRIIa binding antibody compared to P15. Regarding Ab function, P30 had higher ADCP activity relative to both P15 and NP15.

Together, these results show a more robust response in terms of binding and functional Ab features in the non-pregnant as compared to the pregnant women immunized with the 15μg dose of the H1N1 pdm09 vaccine, and in pregnant women who received the 30μg as compared to those who received only 15μg. Notably, among the women vaccinated during pregnancy, those immunized with the 30μg dose had higher increases in NA-specific IgG and NA-specific FcR binding Ab.

### Durability of vaccine-induced antibodies

Given the need for sustained levels of circulating Ab that can be transferred through the placenta to the infant, we first examined the durability of vaccine-induced immunity in maternal serum at delivery as compared to 21 days post-vaccination (**Figure 4, Supplemental Figure 2A**). The number of days between vaccination and delivery was similarly distributed between P15 (median = 120) and P30 (median = 95) (Kruskal-Wallis p = 0.08). Most Ab features declined at the time of delivery, regardless of vaccine dose (**Supplemental Figure 2A**).

**Figure 4.**
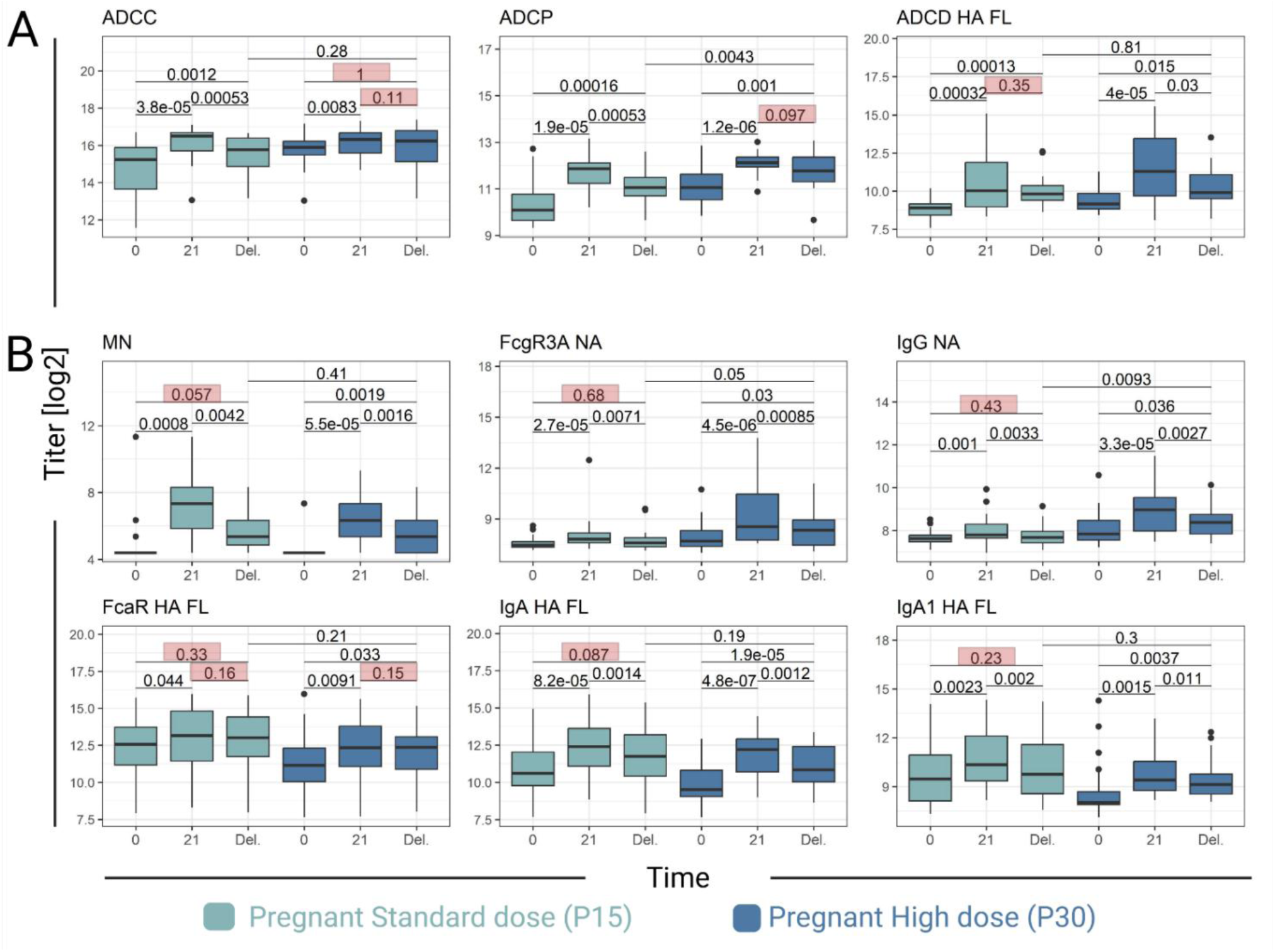
Durability of antibodies elicited by the Novartis H1N1 pdm09 IIV in pregnant women. **(A)** Vaccine-induced features significantly declined from day 21 to delivery (Del.) in only one pregnant cohort but stable in the other. **(B)** Features that significantly declined from Day 21 to Del. in both cohorts, but remained significantly higher at Del. from baseline in only one pregnant cohort. P-values represent paired Wilcoxon for intracohort comparisons and unpaired Wilcoxon for comparisons between P15 vs. P30 at delivery. Non-significant values between time points highlighted in red boxes. Boxplots represent the median, interquartile range, and outliers of the cohort at each time point.

To determine differences in durability between the two cohorts, we identified features that significantly declined at delivery in one cohort but remained stable in the other (**Figure 4A).** HA-specific ADCD had a significant decline at delivery in P30, but not in P15. Alternatively, ADCC and ADCP remained stable from 21 days post-vaccination only in the P30 cohort, whereas P15 showed a significant decline.

Next, we assessed durability based on features that sustained levels above baseline (**Figure 4B).** MN, NA-specific IgG and FcγRIIIa-binding, and HA-specific IgA and IgA1, all showed significant declines in both cohorts from 21 days post-vaccination to delivery. Despite this, the P30 cohort continued to have significantly higher Ab levels at delivery for all of these features at baseline, whereas the P15 cohort declined to baseline levels. HA-specific FcαR-binding had a non-significant decline between 21 days post-vaccination and delivery in both cohorts, but the P30 group remained elevated above baseline.

### Differential transfer and impact of dose on cord blood Ab

We next investigated the differential and dose-dependent transfer of vaccine-induced Ab features by comparing their magnitude in maternal and cord blood within P15 and P30 **(Figure 5)**. Most of the measured Ab features were increased in cord blood as compared to maternal blood, both in the P15 (28/38, 73.7%) and P30 (28/38, 73.7%) groups.

**Figure 5.**
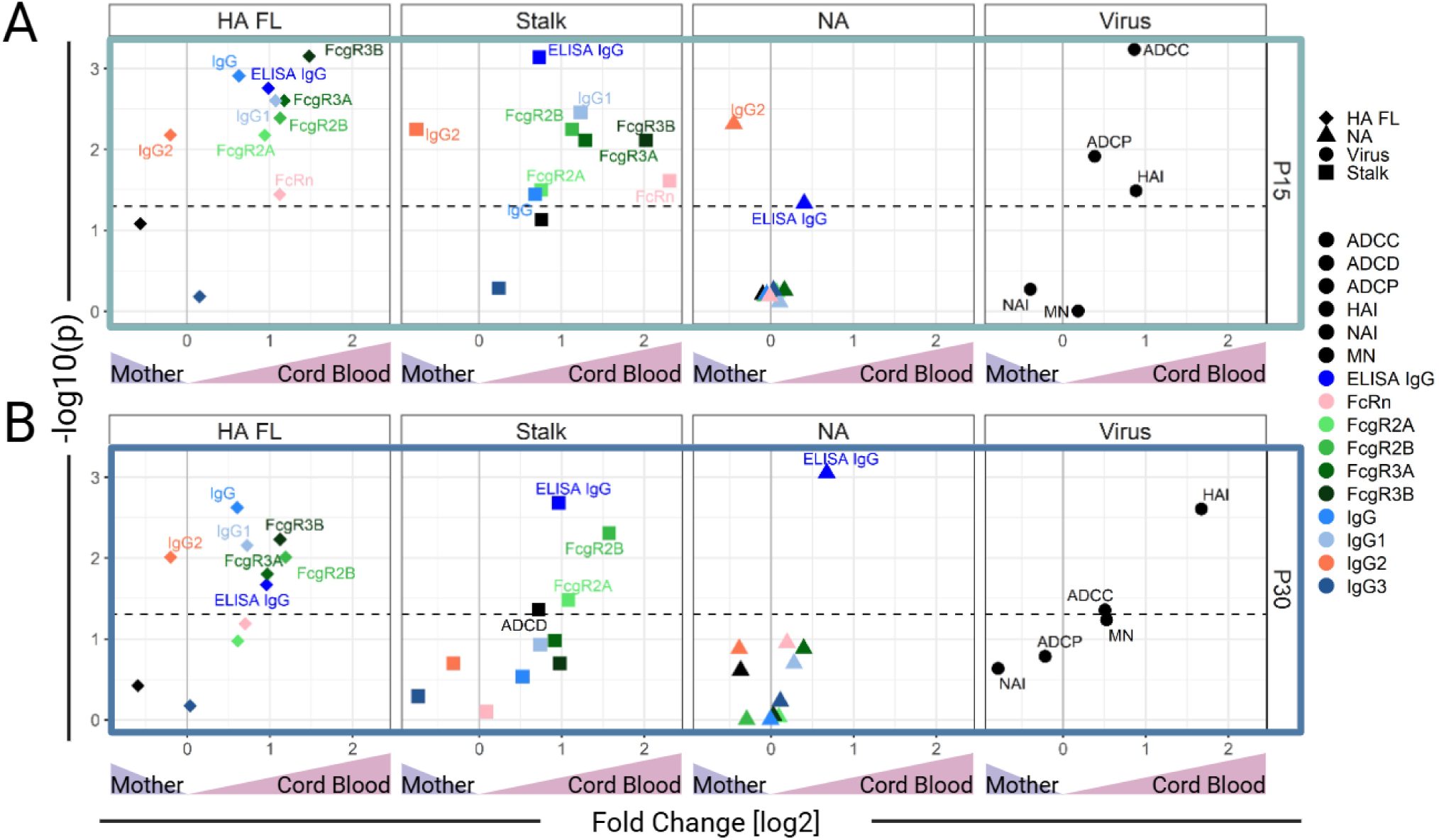
Binding and functional features of antibodies elicited by the Novartis H1N1 pdm09 IIV and transferred to cord blood. Volcano plots depict p-values (paired Wilcoxon test) on the y-axis and differences in fold-change (x-axis) between cord blood and maternal blood at delivery in pregnant women receiving (**A**) 15μg (P15) or (**B**) 30μg (P30). Antigen specificity (full-length hemagglutinin [HA FL], HA stalk domain [Stalk], neuraminidase [NA], and virus) is indicated by shape and separated by column, and features are indicated by color. Horizontal dotted line represents a p-value = 0.05. The solid gray line represents no fold-change; values above 0 had higher expression in cord blood.

Consistent with the maternal responses post-vaccination, there were more HA-specific Ab features increased in cord blood as compared to other antigens, regardless of vaccine dose. HA Ab features increased in cord blood from P15 and P30 groups, including IgG (measured by ELISA and multiplex binding assay), IgG1, FcγRIIIb, FcγRIIIa, and FcγRIIb binding Ab. HA-Stalk-specific IgG (measured by ELISA), FcγRIIb, and FcγRIIa were increased in cord blood from P15 and P30. P15 had a greater number of HA-Stalk-specific features that were higher in cord blood (P15 n=8, P30 n=4). NA-specific IgG was increased in cord blood in both P15 and P30. In contrast, HA-specific IgG2 levels were significantly decreased in cord blood in both P15 and P30 as compared to maternal blood. NA-specific and HA-stalk-specific IgG2 were also lower in the cord blood of P15. HAI and ADCC activity were significantly increased in cord blood in both groups as compared to maternal blood.

### A higher vaccine dose improved NA Ab that persisted through delivery and were transferred to cord blood

To identify features of the Ab response with power to discriminate between pregnant women given high vs. low vaccine dose during pregnancy, the complete serological data measured in maternal blood at day 21 post-vaccination (**Figure 6A**), at delivery (**Figure 6B**), and in cord blood (**Figure 6C**) were examined using a partial least squares discriminant analysis (PLSDA). The leading discriminatory variable (LV1) Ab features contributing to the differentiation of P15 and P30 were ranked in order of importance. NA-specific immunity: NA-IgG, NA-specific FcγRIIIa, and ADCP activity emerged as the most prominent discriminatory factors at day 21 post-vaccination, at delivery, and in cord blood.

**Figure 6.**
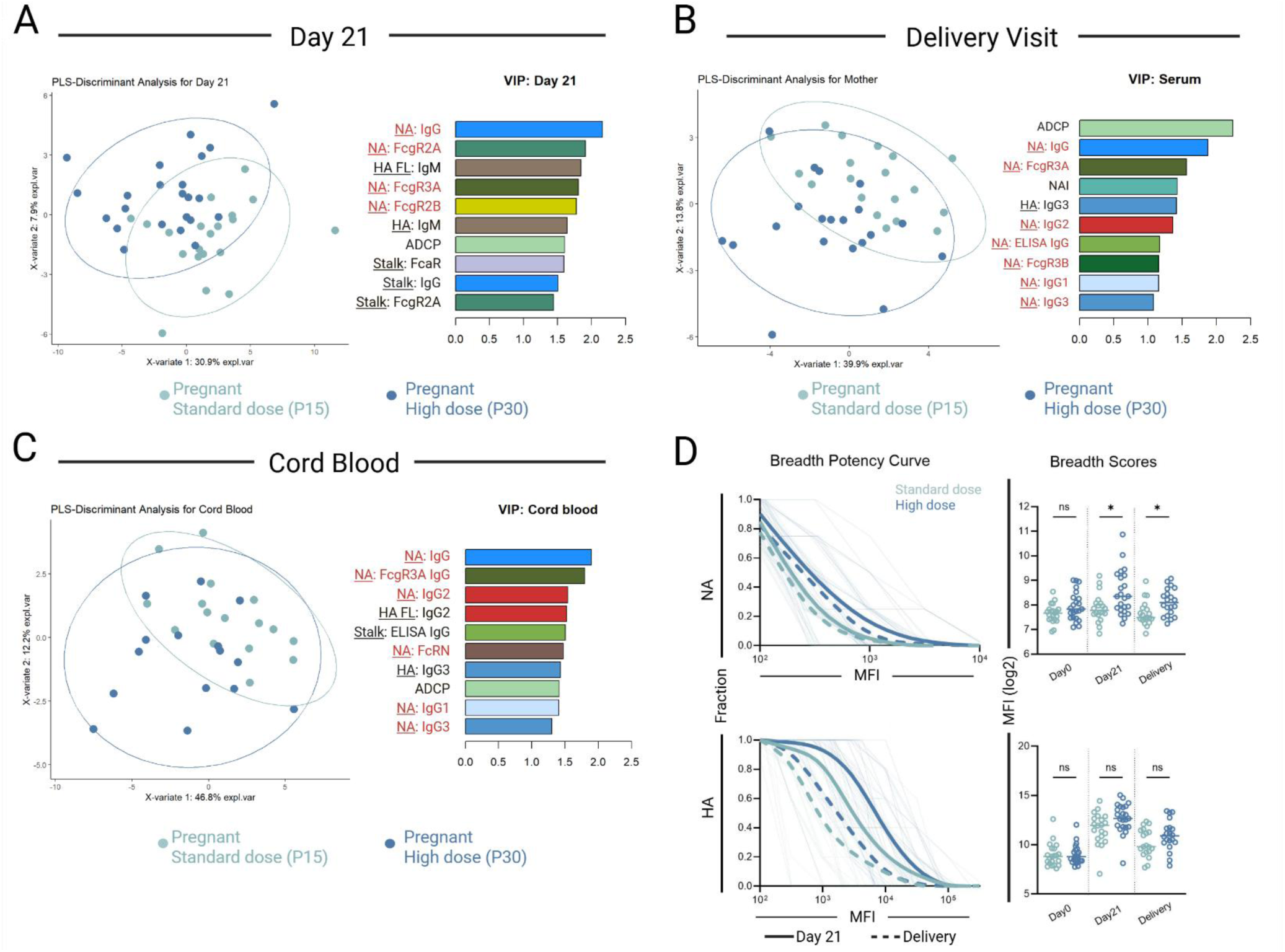
Increased dose of Novartis H1N1 pdm09 IIV improves NA immunity in the mother-infant dyad. (A-C) Partial least squares (PLS) discriminant analysis separating individuals based on exhibited antibody features measured **(A)** in maternal blood at day 21, **(B)** maternal blood at time of delivery, and **(C)** cord blood. Each point represents an individual, and the ellipses represent 95% confidence for each group. (Left) Explanatory variables (X-variate) describe combinations of humoral features, and scores across XVs indicate the percentage of separation described by that axis. (Right) Variable Importance in Project (VIP) scores represent the contribution each feature makes to the PLSDA model, and the top ten are listed. NA-specific features are denoted in red. **D.** (Left) Breadth-potency curves representing the proportion of subjects/antigens with a given level of NA-(top row) and HA-(bottom row) specific IgG, considering NA and HA from multiple influenza strains at day 21 and delivery. Population means are shown with a thick line, and individual subjects are depicted with thin lines. Solid lines depict 21 days post-vaccination, and dashed lines depict delivery. (Right) Breadth scores for each subject. Data represents log-transformed individual data points, and the bar represents the mean. Binding data were compared using Wilcoxon signed-rank test. ∗ p < 0.05, ns = not significant (p > 0.05).

We next evaluated the extent of NA-IgG binding across different heterologous strains of influenza virus H1 or N1 (Supplementary Table 1) by calculating breadth-potency curves and scores, defined as the geometric mean IgG reactivity against NA from different influenza strains (**Figure 6D)**. The same analysis was done for HA. Broad HA-IgG binding was observed post-vaccination; although the mean breadth score was higher in P30 at day 21 and delivery, the difference did not reach statistical significance. In contrast, for NA-IgG binding, the group differences observed at peak immunogenicity, but not at baseline, persisted through delivery.

## Discussion

To interrogate pregnancy-associated changes in vaccine responses, we leveraged archived samples from a clinical study that evaluated the safety and immunogenicity of the Novartis inactivated influenza A (H1N1) pdm09 vaccine in pregnant and non-pregnant women. The trial design included a dosage level comparison in pregnant women, i.e., 15μg vs. 30μg, which allowed us to test the hypothesis that maternal-infant immunity can be strengthened by increasing the vaccine dose.

Our study is unique from previous reports describing pregnancy-associated changes in influenza vaccine responses in that it explored an H1N1 pandemic strain to which mothers and infants are highly susceptible, examined a wide array of binding and functional Ab features specific for key viral antigens, i.e. HA, HA stalk, and NA, and monitored vaccine-induced Ab longitudinally, before and after vaccination and up to the time of delivery, and in the infants at birth (cord blood).

The H1N1 pdm09 vaccine exhibited a favorable safety profile and adequate immunogenicity. Despite most participants exhibiting HAI seroconversion and seroprotection post-vaccination, a lower proportion of pregnant women in the standard 15μg group met these marks, as compared to either the non-pregnant immunized with the same dose or the pregnant group immunized with the 30μg dose. Pregnant women receiving the 15μg dose exhibited a more subdued response to the vaccination, as indicated by the proportion of increased Ab features (Ab binding and Fc-mediated function) post-vaccination across all antigens. The doubling of the vaccine dose in the pregnant group had a positive effect, as it expanded Ab responses broadly post-vaccination, with more Ab features increasing. The 30μg dose elicited stronger NA-specific immunity, specifically IgG and Fc receptor binding Ab, as well as NA-reactive Ab capable of engaging innate cell function, i.e., ADCP. Notably, the superiority of the 30μg dose was evident by the persistence of NA-specific IgG in maternal blood up to the time of delivery, as well as the more abundant NA-IgG and HAI in the infants at birth. The NA-specific features emerged as the main discriminators of maternal serological profiles based on vaccine dose 21 days after vaccination and at delivery, and among the top dose-discriminatory features in cord blood.

NA is the second most abundant glycoprotein on the surface of Influenza A viruses and has important roles in the virus life cycle and pathogenesis, facilitating the egress of new virions from the host cell and allowing the virus to traverse the mucus layer and cleave sialic acid on the surface of target cells (22). NA-specific Ab are known to have a protective role in vivo(23–27). In controlled human infection studies with H1N1 Influenza viruses, NA Ab were inversely correlated with the severity of disease (28). They have also been associated with reduced virus shedding and illness in infected adults in community settings (27, 29). Antibodies that inhibit NA activity (NAI) have been identified as an independent correlate of protection (24, 30, 31), and as NA exhibits lower antigenic variation (32), NA immunity could be more effective against strain seasonality. Hence, enriching NA-specific Abs through antenatal vaccination may be an appealing approach to bolster vaccine effectiveness in the maternal-infant dyad and beyond.

The CDC and ACIP recommend the use of influenza vaccines containing four times the standard dose in adults 65 years of age or older, and this approach has proven to be more efficacious in this population (20). An inactivated high-dosage (60μg of HA per strain) seasonal influenza vaccine was reported to elicit eight times higher NAI Ab than the standard vaccine among elderly subjects (33). While the optimal anti-NA Ab responses and the NA necessary for inducing such responses are uncertain, a greater NA dosage than that contained in the standard 15μg vaccine may be beneficial. The amount of NA in seasonal influenza vaccines is not regulated, nor do vaccine manufacturers report it, though it is known to vary among formulations (34). Compared to HA antigenicity, NA antigenicity is meagerly characterized and documented. NA Ab are not routinely measured in clinical studies, and the extent to which seasonal vaccines elicit these Ab is uncertain. The H1N1 pdm09 monovalent vaccine used in the DMID 09-0072 study described here had the highest content of NA among several lots tested (35).

Prior studies investigated the safety and immunogenicity of varying doses of H1N1 pdm09 during pregnancy. One of them compared the safety and immunogenicity of monovalent H1N1pnd09 administered twice, 21 days apart, in pregnant women at two dosage levels: 25μg and 49μg (36). It was reported that a single 25μg dose was highly immunogenic, achieving >90% seroprotection with no apparent benefits from a second dose. Consistent with our findings, HAI titers were found to decline from post-vaccination levels to delivery, although still found in cord blood at higher levels as compared to those of maternal blood. A subsequent study involved the administration of H1N1pnd09 adjuvanted with MF59 or unadjuvanted to pregnant women in doses ranging from 3.75-15μg, while a non-pregnant control group received 7.5μg of MF59-adjuvanted H1N1pnd09 (37). A higher HAI response was observed in the non-pregnant group compared to pregnant women immunized with the same dosage of the adjuvanted vaccine. The study design did not include 15μg unadjuvanted vaccination in both groups and could not assess the adequacy of this dose in pregnancy. We are not aware of any other study that comparatively evaluated influenza vaccination (H1N1pdm09) in both pregnant and non-pregnant women, including both the currently approved standard (15μg) and a twice as high vaccine dose.

Among studies that examined H1N1 pdm09 immunogenicity in other age groups, one reported a higher HAI fold rise in adults immunized with a 30μg dose compared to the standard 15μg dose (38). In their discussion, the authors still favored the 15μg dose based on high seroconversion achieved and practical considerations of vaccine availability in a pandemic setting, although they acknowledged the need for trials in other populations (38). A significant limitation of the studies discussed above is the reliance only on HAI data. It is clear from our results that a more complete and nuanced landscape of influenza immunity emerges from a deeper analysis of Ab features. Although HAI is used as an indicator of protection, NA Ab might be equally or more important in preventing disease (39, 40).

There are no preferential vaccine recommendations for immunization of pregnant women against influenza, A recent study compared a single dose of 15μg or 30μg or two doses of 15μg one month apart of a seasonal inactivated influenza vaccine in pregnant women with HIV(41) and reported mothers receiving the single 30μg vaccine had higher seroconversion and seroprotection compared to either other group, and their infants had a higher rate of seroprotection than those receiving a single dose of 15μg. These, along with our results, support the value of a higher dose vaccine to improve maternal immunization against influenza.

A valid question is whether the enhanced responses to a higher dose vaccine impact vaccine potency. Our study did not include disease surveillance, and the lack of influenza illness data prevents us from drawing conclusions on vaccine effectiveness and from evaluating associations between the Ab features measured and the incidence of disease. Another limitation of our study is the relatively small sample size, which reduced our ability to identify statistically significant differences between groups. The H1N1 pdm09 vaccine included only one subtype of HA (H1) and NA (N1); it would be important to confirm the effect of the dose on vaccine responses in a more diverse set of antigens. A technical limitation relates to the functional assays in the multiplex format; the use cell lines instead of freshly isolated human primary cells make them surrogates, rather than true indicators of Ab engagement with innate immune cells.

In summary, our results demonstrated that humoral immune responses in pregnant women immunized with the H1N1 pdm09 vaccine were enhanced by administering twice the standard vaccine dose. Importantly, the 30 μg dose elicited higher levels of NA-specific Ab with broad reactivity across influenza strains, which persisted through delivery and remained higher in cord blood. The potential for improving maternal and infant influenza immunity by increasing the recommended vaccine dose during pregnancy warrants further investigation. Studies of larger sample sizes, with diverse populations and longitudinal follow-up, will be particularly relevant to inform public health decisions.

Our findings also emphasize the need to consider pregnant women and their infants in preparedness for emergent threats.

## Materials and Methods

### Clinical samples

Serum samples from DMID study 09-0072 were provided by the National Institute of Health (NIH) Division of Microbiology and Infectious Diseases (DMID). These specimens were stored at -80°C at Fisher Bioservices. Blood was obtained at baseline and at day 21 post-vaccination. For participants who were pregnant, blood was also obtained at delivery and from cord blood. All participants were HIV, HBV, and HCV negative, and were provided with written informed consent before enrollment. The biomarker analysis was approved by UMB and Dartmouth Institutional Review Boards under the non-human subject research category.

### HAI, MN, and NAI measurements

Sera pre-treated with receptor-destroying enzyme (RDE, Denka Co., New York, NY) were two-fold serially diluted and tested for HAI using four HA units of H1N1 pdm09 A/California/07/2009 in phosphate-buffered saline (PBS, pH 7.4) and 0.5% turkey red blood cells, as previously described (42, 43). HAI titers were reported as the reciprocal of the highest serum dilution that resulted in complete inhibition of hemagglutination. Samples for which no inhibition was observed at the initial 1:10 dilution were assigned a titer of 5.

MN titers were determined using a live virus-based ELISA, as previously described (44). In brief, RDE-treated sera were two-fold serially diluted and incubated with 100 50% tissue culture infectious dose (TCID_50_) of H1N1 pdm09 A/California/07/2009 for 1h at room temperature. The mixtures were added to MDCK-SIAT1 cells (Sigma, St. Louis, MO) and incubated overnight at 37°C, 5% CO_2_. Infected cells were detected using mAb specific to influenza A nucleoprotein (Millipore Sigma, Burlington, MA), followed by peroxidase-labeled goat anti-mouse IgG diluted 1:2,000 (Seracare, Milford, MA). MN titers were reported as the reciprocal of the highest serum dilution that yielded >50% reduction in Optical Density (OD) value as compared to wells containing only the virus. Samples for which no neutralization was observed at the initial 1:40 dilution were assigned a titer of 20.

NAI titers were determined using a previously reported enzyme-linked lectin assay (26) with modifications. Briefly, microtiter plates were coated with 75µg/mL of fetuin (Sigma-Aldrich) overnight at 2-8°C. Serum samples were two-fold serially diluted and mixed with an equal volume of a recombinant virus expressing N1 of A/California/07/2009 and a chimeric H6 gene. The mixtures were then incubated in fetuin-coated plates for 16-18 h at 37°C. Plates were washed, and 1 µg/mL of peroxidase-labeled peanut agglutinin (Sigma-Aldrich) was added, followed by a 2-hour incubation. N1 signals were probed with O-phenylenediamine dihydrochloride (Thermo Fisher Scientific, Waltham, MA), and the colorimetric reaction was stopped by adding 1 N H_2_SO_4_. OD_490 nm_ values were measured using a Victor V plate reader. NAI titers were determined as the reciprocal of the highest serum dilution that yielded >50% inhibition of NA activity. A titer of 5 was assigned to samples for which <50% inhibition was observed at the initial 1:10 dilution.

### HA and NA ELISA

Immulon 2HB 96-well plates were coated with either 1µg/ml of recombinant HA or HA stalk based on an assay format previously described (45), or with 2µg/ml of recombinant NA from A/California/07/2009 and incubated overnight at 2-8°C (46). Following incubation, plates were washed and blocked with 5% non-fat dry milk (NFDM; Nestlé, Vevey, Switzerland) in PBS containing 0.05% Tween-20 for 1 h at room temperature (22°C). After washing, diluted samples and controls were added to the plates and incubated at room temperature for 2 h. After washing, bound Ab were detected using HRP-labeled goat anti-human IgG diluted 1:5,000 (Seracare), followed by TMB substrate. Color development was stopped by adding 1M Phosphoric Acid.

### Fc Array Assay

Serum antibodies specific for a panel of antigens (**Supplemental Table 1**) were characterized using the Fc multiplexed array, as described previously (47, 48). Briefly, antigens were covalently conjugated to fluorescently coded magnetic beads (MagPlex Microspheres, Luminex Corporation, Austin, TX). Serum dilutions, appropriate for each detection reagent varying from 1:250 to 1:10,000, were prepared in Assay Wash Buffer (AWB) and added to washed beads before incubating for 2 h. Following binding and washing, antigen-specific antibodies were detected by R-phycoerythrin (PE)-conjugated secondary reagents specific to human immunoglobulin isotypes and subclasses or site-specifically tetramerized Fc receptors (49). Following standard incubation and washing processes, beads were resuspended in AWB (Luminex Corporation), and Median Fluorescent Intensity (MFI) values were acquired on a FlexMap 3D array reader (Luminex Corporation). Pooled human polyclonal serum IgG IVIG (Octagam 5%, Pfizer, New York, NY) was used as a positive control, and PBS-TBN (0.1% BSA, 0.02% sodium azide, 150 mM sodium chloride, 50 mM sodium phosphate monobasic anhydrous, 0.05% Tween-20, pH 7.4) buffer was employed as a negative control. The data reported for each sample represent the mean of duplicates.

### ADCC and ADCP

A luciferase reporter cell-based FcγRIIIa activation assay and a FcγRIIa activation assay were used to measure ADCC and ADCP functions, respectively (50). Briefly, MDCK-SIAT1 cells were pre-seeded at 10^4^ cells/well in 96-well white flat-bottom plates (Costar) overnight and were infected with H1N1pdm09 A/California/07/2009 at a multiplicity of infection (MOI) of 5 at 37°C, 5% CO_2_ for six hours. RDE-treated sera (25 µL/well) were added along with either FcγRIIIa(V158)-expressing reporter cells (5X10^4^/25 µL/well) (Promega, Madison, WI) for ADCC or FcγRIIa (H131)-expressing reporter cells (Promega, 5X10^4^/25 µL/well) for ADCP to infected cells. After further incubation at 37°C and 5% CO_2_ overnight, 50 µL of Bio-Glo luciferase assay reagent (Promega) was added to the wells, and luminescence in Relative Light Units (RLU) was measured 15 minutes later using a Victor V multilabel reader. ADCC or ADCP activities were expressed as the RLU at serum dilution of 1:20,000 or 1:2000, respectively.

### ADCD

ADCD was measured as previously described (51). Briefly, heat-inactivated sera were incubated with antigen-conjugated multiplex assay beads on a shaker for 2 h at room temperature as described above. Optimal serum dilutions (falling within linear detection range) were determined in a pilot experiment that tested a subset of samples over a range of dilutions (data not shown). Guinea pig complement serum (Cedarlane Labs, Burlington, NC) was diluted 1:60 in gelatin veronal buffer (Sigma-Aldrich) and mixed with diluted serum samples at 37°C with shaking for 20 minutes. Following this incubation, cold EDTA solution (15 mM in 1XPBS) was added to prevent further complement activation. After washing, samples were incubated with biotin-conjugated goat anti-C3b (1µg/mL) (Icllab, Portland) at room temperature for one hour, followed by staining with Streptavidin-R-Phycoerythrin (Millipore Sigma) reagent per the manufacturer’s recommendation. A final wash was performed, samples were resuspended in Luminex sheath fluid, and MFI was acquired on a FlexMap 3D reader (Luminex). Influenza A virus control serum negative (IBL-America 4112) with heat-inactivated complement serum and assay wash buffer (PBST) were used as negative controls. IVIG (Pfizer Octagam 5%) was used as a positive control. Samples were tested in replicates, and results were averaged.

### Data Analysis

Data were analyzed using GraphPad Prism software (V9.0 and 10.0) and the statistical platform The R Project for Statistical Computing (V4.4.0). First, raw measurements were compared with the negative/positive control data. Measurements were logarithmically transformed before statistical analysis was performed. In particular, the *stats* package (V4.4.0) and the *mixOmics* package (V6.28.0) were used to analyze the data; the *ggplots* package (V3.5.1) and the *ggrepel* package (V0.9.5) of the platform were used to generate the boxplots, volcano, PLSDA, and VIP plots. Statistical significance was calculated using Wilcoxon test or unpaired student’s T-test. Breadth–potency curves were defined as the proportion of subjects exhibiting a signal above a given intensity for antigen specificities. Curves were generated using the LOWESS curve fit method in Prism for each respective subject group. Breadth scores were calculated by taking the geometric mean across antigen specificities for each subject.

## Data Availability

All data produced in the present study are available upon reasonable request to the authors

## Acknowledgments

The DMID 09-0072 Clinical Study Group: Shital Patel, M.D, Baylor College of Medicine; Lisa Jackson, M.D., M.P.H., Group Health Research Institute; Sharon E. Frey, M.D, Saint Louis University; Karen L. Kotloff, MD, University of Maryland, Baltimore, and Kathryn Edwards, M.D, Vanderbilt University

The authors would like to thank DMID scientists Dr. Chris Roberts and Melinda Tibbals for facilitating access to clinical samples, Joshua Weiner for his invaluable guidance and support in the Ab binding analyses, and Haye Nijhuis for production of influenza proteins.

## Author contributions

D.K. and M.K. designed and executed the experiments and analyzed the data. J.Z., M.S.Z., A.H., and P.L.M. performed bioinformatic analysis and visualization. M.E.A., M.F.P., and H.X. conceived the project and supervised laboratory analyses. L.C. provided reagents. G.S. served as a principal clinical investigator for this study. D.K., M.K., P.L.M., H.X., and M.F.P wrote the manuscript with input from co-authors. All authors reviewed and edited the manuscript.

