## Supplementary Files for "Increased dose of H1N1 pandemic influenza vaccine during pregnancy improves immunity in mothers and infants"

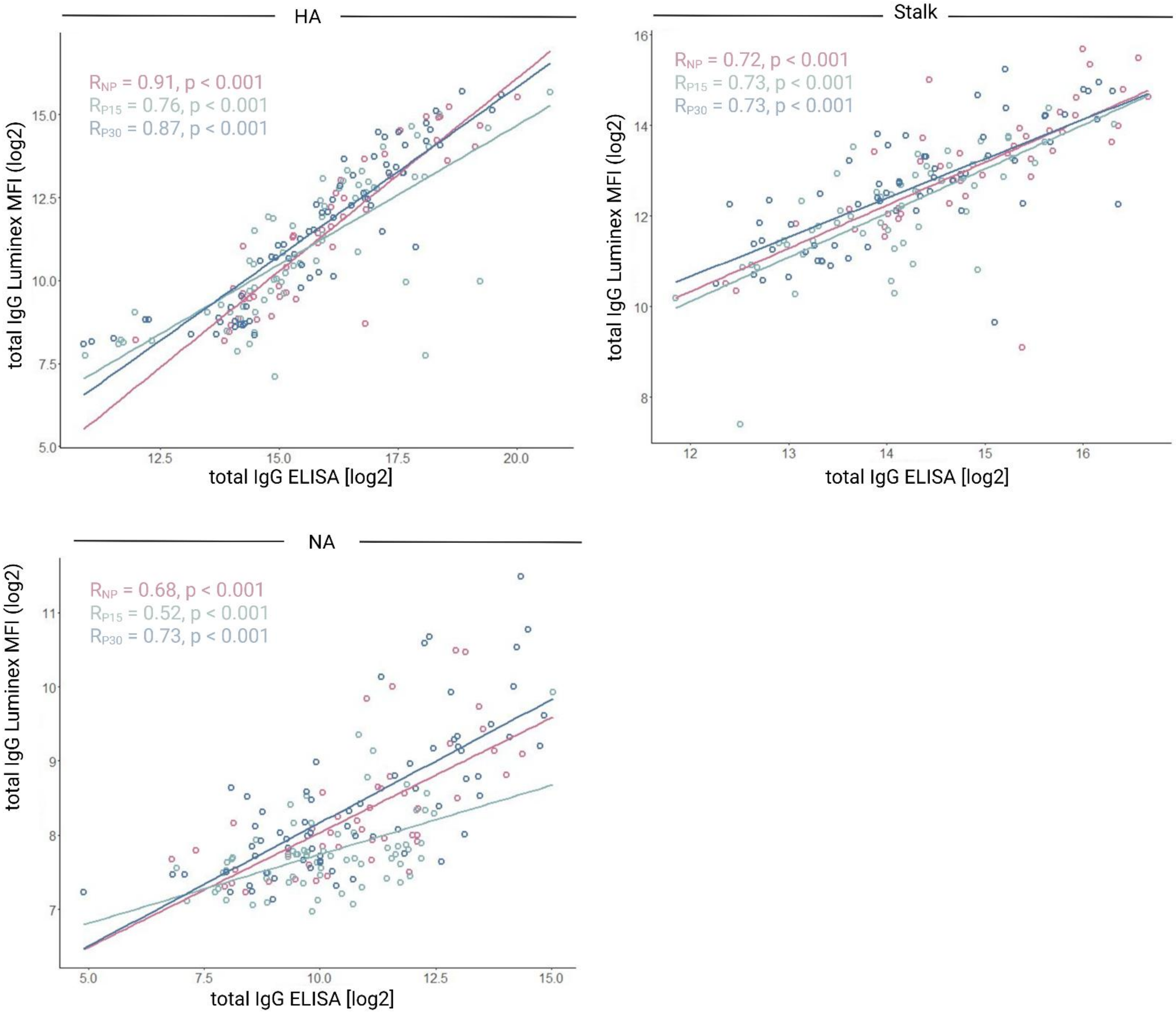


**Supplemental Figure 1. Correlation between IgG from ELISA and Luminex binding assay.**

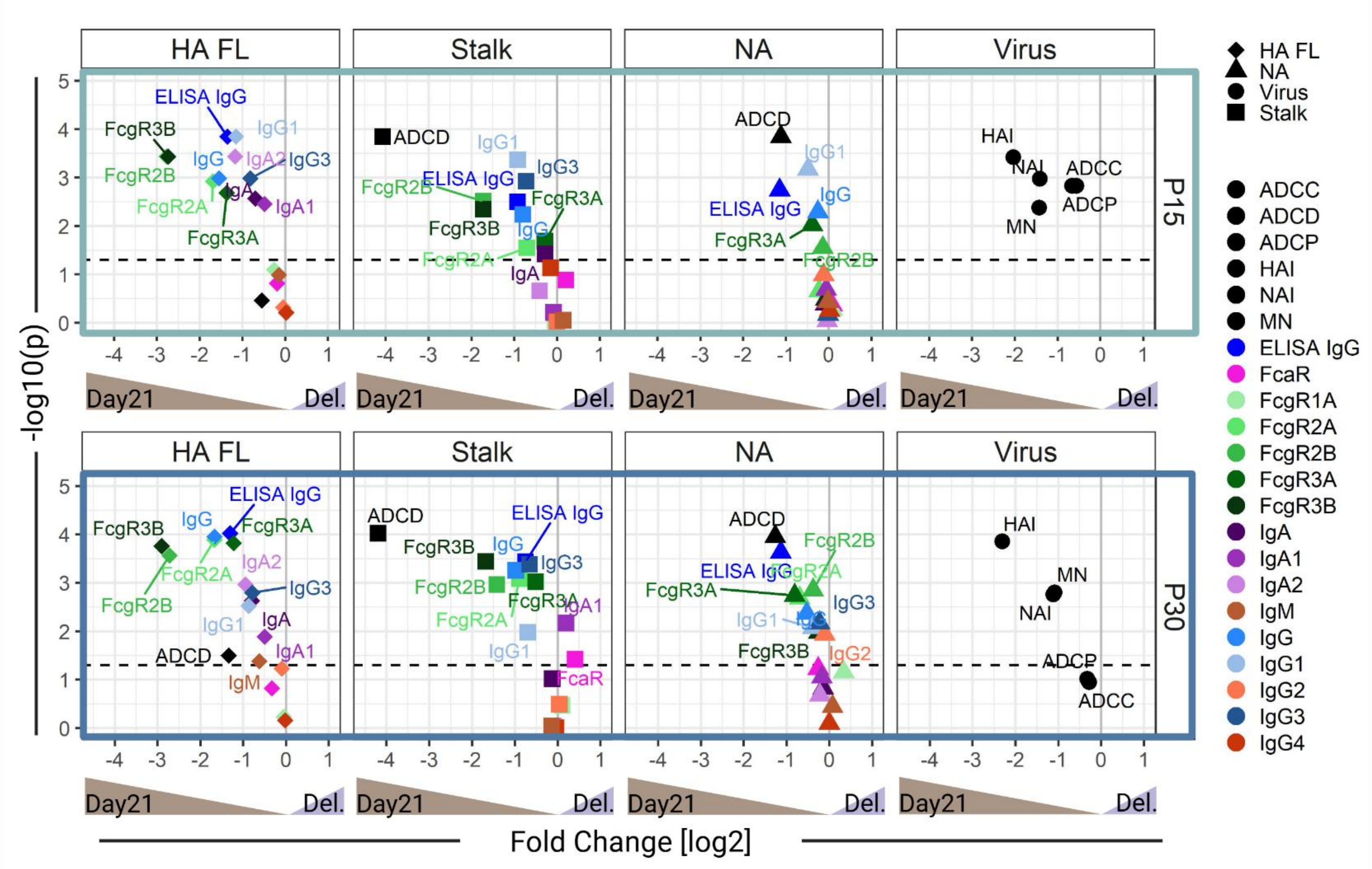


**Supplemental Figure 2.** Volcano plots depicting p-values (paired Wilcoxon test) and fold-changes in titers at 21 days post-vaccination (Day21) vs. at delivery (Del.) in pregnant women receiving 15µg (P15) or 30µg (P30) of the Novartis H1N1 pdm09 vaccine. Horizontal dashed line represents p = 0.05. Vertical grey line represents no fold change, values above 0 had higher expression at delivery.

**Supplemental Table 1.** List of antigens and sources.

| Antigen | Source | Catalog # |
|---|---|---|
| H1 Headless HA (Stem) based on H1N1 (A/California/04/2009) | UMB | PLC22 14 |
| H1N1_Michigan_NA | Sino | 40568-V07H |
| H1N1_Cali_NA | Sino | 11058-V08B |
| H1N1_M1_PuertoRico | immunetech | it-003-045ep |
| H1N1_Cali_HA | Sino | 11055-V08H2 |
| H1Cal09+Fib (Full-length ectodomain) based on H1N1 (A/California/04/2009) | UMB | PLC22 09 |
| H1N1_WI_HA | Sino | 40787-V08H |
| H1N1_Cali_TM_HA | immunetech | it-003-sw12Δtmp |
| H1N1_Brisbane_HA1 | immunetech | it-003-0012p |
| H1N1_Brisbane_NA | Sino | 40767-V08B |
| H1N1_Michigan_HA | Sino | 40567-V08H1 |
| H1N1_WI_NA | Sino | 40785-V08B |
| H1N1_Brisbane_HA | Sino | 40719-V08H |
| a-IgG | Southern Biotech | 2048-09 |
| a-IgG1 | Southern Biotech | 9054-09 |
| a-IgG2 | Southern Biotech | 9070-09 |
| a-IgG3 | Southern Biotech | 9210-09 |
| a-IgG4 | Southern Biotech | 9200-09 |
| a-IgA | Southern Biotech | 2050-09 |
| IgA2 | Southern Biotech | 9140-09 |
| IgA1 | Southern Biotech | 9130-09 |
| a-IgM | Southern Biotech | 9020-09 |
| FcγR2aR131 | In House | |
| FcγR2b | In House | |
| FcγR3aV158 | In House | |
| FcγR3bNA2 | In House | |
| FcαR | In House | |
| FcRn | In House | |
| FcγR1 | In House | |
